# Histopathology and Ultrastructural Findings of Fatal COVID-19 Infections

**DOI:** 10.1101/2020.04.17.20058545

**Authors:** Benjamin T. Bradley, Heather Maioli, Robert Johnston, Irfan Chaudhry, Susan L. Fink, Haodong Xu, Behzad Najafian, Desiree Marshall, J. Matthew Lacy, Timothy Williams, Nicole Yarid

**Author notes:** Corresponding author: Dr. Benjamin T. Bradley MD/PhD, 1959 NE Pacific St, Box 357470, Seattle, WA 98195. These authors contributed equally (Benjamin T. Bradley, Heather Maioli). IRB approval for this study was waived by the University of Washington.

## Abstract

**Background:** SARS-CoV-2 is the cause of an ongoing pandemic with a projected 100,000 to 240,000 U.S. deaths. To date, documentation of histopathologic features in fatal cases of COVID-19 has been limited due to small sample size and incomplete organ sampling.

**Methods:** Post-mortem examinations were performed on 12 fatal COVID-19 cases in Washington State during February-March 2020. Clinical and laboratory data were reviewed. Tissue examination of all major organs was performed by light microscopy and electron microscopy. The presence of viral RNA in sampled tissues was tested by RT-PCR.

**Results:** All 12 patients were older with significant preexisting comorbidities. The major pulmonary finding was diffuse alveolar damage in the acute and/or organizing phases with virus identified in type I and II pneumocytes by electron microscopy. The kidney demonstrated viral particles in the tubular epithelium, endothelium, and podocytes without significant inflammation. Viral particles were also observed in the trachea and large intestines. SARS-CoV-2 RNA was detected in the cardiac tissue of a patient with lymphocytic myocarditis. RT-PCR also detected viral RNA in the subcarinal lymph nodes, liver, spleen, and large intestines.

**Conclusion:** SARS-CoV-2 represents the third novel coronavirus to cause widespread human disease since 2002. Similar to SARS and MERS, the primary pathology was diffuse alveolar damage with virus located in the pneumocytes. However, other major organs including the heart and kidneys may be susceptible to viral replication and damage leading to increased mortality in those with disseminated disease. Understanding the pathology of SARS-CoV-2 will be essential to design effective therapies.

## Introduction

In December 2019, a novel coronavirus (SARS-CoV-2) was identified in Wuhan, China from a cluster of severe pneumonia cases.(1) The virus and the disease it causes (COVID-19) have now spread globally and are responsible for the first pandemic since the 2009 H1N1 influenza virus. During the four months following its emergence, the community of healthcare workers and researchers have acted quickly to sequence the virus, establish transmission chains, elucidate the receptor, and test therapeutics.(2)(3)

These efforts have revealed similarities between SARS-CoV-2 and the related virus, SARS-CoV. Both have similar clinical presentations with the highest viral load identified in lower respiratory samples.(4) SARS-CoV-2 RNA has also been detected from stool and blood samples prompting concerns for multiorgan involvement which would mimic observations in SARS.(5) The viruses also share a common cellular entry receptor, angiotensin converting enzyme 2 (ACE2).(3) In contrast, SARS-CoV was responsible for a limited disease outbreak with high mortality whereas SARS-CoV-2 has caused a greater number of infections with relatively lower mortality.(6)

Despite early successes in characterizing the virus, reports on the histopathologic manifestations and tissue tropism of fatal COVID-19 cases remain sparse.(7,8) This absence of information reflects the acute and overwhelming number of cases straining medical systems globally. Accurate documentation of the pathobiology in fatal COVID-19 infections may have implications on patient management and antiviral development. Differences noted between SARS-CoV and SARS-CoV-2 may suggest novel mechanisms to target.

Available histopathologic studies have demonstrated pulmonary and gastrointestinal involvement by SARS-CoV-2 but are limited by small case numbers or limited sampling adequacy.(7–9) This study addresses those shortcomings by documenting a series of 12 fatal COVID-19 cases which occurred in Washington State during February-March 2020. Systematic histopathologic evaluation of all major organs was performed by a combination of light microscopy, electron microscopy, and molecular detection.

## Methods

### Patient Selection and Autopsy Procedure

IRB approval for this study was waived by the University of Washington. Patients with a positive antemortem or post-mortem SARS-CoV-2 result were considered eligible for enrollment. Preliminary testing for SARS-CoV-2 was performed at the Washington State Department of Health (WSDOH) Public Health laboratory. Confirmatory testing was done by the Centers of Disease Control (CDC) in Atlanta, Georgia. Both locations used the CDC-designed 2019 nCoV real time RT-PCR assay for viral detection.(10) Autopsies were performed at the King County Medical Examiner’s Office and Snohomish County Medical Examiner’s Office in negative-pressure isolation suites. Given safety concerns, an in-situ dissection was performed for seven cases. The remaining five were examined by standard autopsy procedure. In one case fresh tissue was collected for electron microscopy RT-PCR studies.

### Histologic Examination

Autopsy material was fixed in 10% neutral buffered formalin and submitted for standard processing with hematoxylin and eosin (H&E) staining. Evaluation of H&E sections was performed by consensus agreement of four board-certified forensic pathologists (N.Y., T.W., J.M.L, D.M.) with expert guidance provided in cardiothoracic pathology (H.X.) and renal pathology (B.N.).

### Ultrastructural Examination

Samples were placed in 1/2 strength Karnovsky fixative. Tissue was then post fixed in 1% osmium tetroxide, processed following standard transmission electron microscopy procedures and embedded in PolyBed 812. Suitable sections were identified by toluidine blue staining. Thin sections were examined using a Tecnai G2 Spirit Bio-Twin transmission electron microscope and digital images and measurements were acquired using the AMT image capture software.

### Molecular Detection of Viral RNA in Tissue

RNA was extracted from 0.5 µg of tissue using the Direct-zol RNA Miniprep Plus kit (Zymo Research, Irvine, CA, USA). The CDC 2019-Novel Coronavirus Real-time RT-PCR Diagnostic Panel assay was adapted to a two-step procedure using the 2019 nCoV N1 and N2 primer/probe sets (IDT, Coralville, Iowa, USA).(10). Complementary DNA was synthesized using the iScript cDNA kit (Bio-Rad). Samples were tested twice in duplicate using iTAq Universal Probes Supermix (BioRad). Ct values less than 40 were considered positive.

## Results

### Demographics and Clinical Characteristics

The average age of our cohort was 70.4 years (range 42 to 84 years old). Seven members of the cohort were part of a single cluster from a long-term care facility.(11) All patients had significant comorbidities, with the most common being hypertension, chronic kidney disease, obstructive sleep apnea, and metabolic disease including diabetes and obesity (Tables 1 and S1).

**Table 1:** Fatal COVID-19 Infections: Patient Characteristics, Comorbidities, Symptoms, Radiographic findings, and Select Initial Laboratory Values.

| Case Number | Initial symptoms | Time from symptom onset to admission (Days) | Time from symptom onset until intubation (Days) | Time from symptom onset to death (Days) | Radiographic findings | Elevated Creatinine | Elevated Troponin | Secondary Bacterial Infection |
| --- | --- | --- | --- | --- | --- | --- | --- | --- |
| 1 | Cough, fever, chills, loose stool, fatigue, respiratory distress | 4 | 4 | 10 | Bilateral multifocal patchy airspace opacities | + | - | - |
| 2 | Acute renal failure, AMS, cough, acute cardiomyopathy, acute respiratory distress | 2 | 2 | 2 | Increased PA and interstitial markings | + | - | - |
| 3 | 40 C fever, respiratory distress, tachycardia | 1 | Not applicable | 2 | Bilateral patchy opacities | - | - | - |
| 4 | Cough, tactile fever, body aches, respiratory distress | 1 | 1 | 1 | Bilateral diffuse scattered opacities | - | - | - |
| 5 | Cough, respiratory distress, fever | 5 | 5 | 13 | Widespread bilateral opacities | - | - | - |
| 6 | Respiratory distress, AMS | 1 | Not applicable | 2 | Bibasilar atelectasis or consolidations with small pleural effusions | - | - | - |
| 7 | Acute respiratory distress, cough | 7 | Not applicable | 13 | Multifocal bilateral opacities | + | - | *Pseudomonas aeruginosa* |
| 8 | Respiratory distress, hypotension, tachycardia, fever | 3 | 3 | 7 | Multifocal bilateral opacities | + | + | MSSA and Influenza A |
| 9 | AMS, respiratory distress, fever | 3 | Not applicable | 12 | Bilateral interstitial opacities and asymmetric edema on the right | - | - | - |
| 10 | Respiratory distress, AMS, hypotension | 1 | Not applicable | 1 | Complete opacification of the left hemithorax with additional right lower lobe and middle lobe opacities | + | - | - |
| 11 | Fever, cough, nausea and vomiting | 1 | 5 | 7 | Bilateral multifocal opacities | - | + | - |
| 12 | Fever, headache, leukopenia | 5 days | 12 | 14 | Bilateral multifocal opacities | - | - | - |

Hypertension (HTN), Pulmonary hypertension (pHTN), Chronic kidney disease (CKD), Obstructive sleep apnea (OSA), DM2 (Type-2 Diabetes), Altered mental status (AMS), Atrial fibrillation (AFib), Congestive Heart Failure (CHF), Heart failure with preserved ejection fraction (HFpEF), Chronic Obstructive Pulmonary Disease (COPD), Aortic Stenosis (AS), Hyperlipidemia (HLD), deep vein thrombosis (DVT), Mitral regurgitation (MR). Elevated troponins defined by >0.4 ng/mL, elevated creatinine defined by 1.5 mg/dL, fever defined by >37.5C.

The most frequent presenting symptoms were respiratory distress (83.3%), fever (58.3%), and cough (50%). Less commonly encountered initial symptoms included altered mental status and gastrointestinal distress. During the course of hospital admission, five patients experienced acute kidney injury demonstrated by elevated creatinine and two patients developed elevated troponins. Clinically documented pulmonary co-infections were observed in two patients. One case had Influenza A and methicillin-sensitive *Staphylococcus* aureus; the other had sputum cultures positive with *Pseudomonas aeruginosa*. Excluding those who declined resuscitative measures, all patients were intubated. The median time from symptom onset to intubation was 4.6 days with most intubations occurring at the time of admission. Mortality following symptom onset varied from 1 to 14 days with a median time of 7 days. Demographics and patient characteristics are described in Table 1.

### Gross Autopsy Examination

All decedents examined by standard autopsy had heavy, edematous lungs which averaged 1049 grams for the right lung and 755 grams for the left lung. Intraparenchymal hemorrhage was appreciated in one case. There was no evidence of pulmonary consolidation. The volume of pleural fluid was highly variable and ranged from 0-450 mL per pleural space. Two cases had evidence of sub-segmental pulmonary emboli.

One case demonstrated mild splenomegaly (350gm). Scattered punctate subarachnoid hemorrhages were observed in one brain. Additional gross findings demonstrated non-specific chronic changes including hepatic sinusoidal congestion (100%), hypertensive renal surface changes (80%), and varying degrees of atherosclerotic coronary artery disease (60%).

### Histologic Examination

In all cases, sections of lung demonstrated enlarged, reactive type II pneumocytes with nucleomegaly and prominent nucleoli. Changes of acute and/or organizing diffuse alveolar damage (DAD) were present in 75% of cases as evidenced by the presence of intra-alveolar fibrin, hyaline membranes, and loosely organizing connective tissue in the alveolar septal walls (Figure 1). Half of the cases showed focal areas of acute bronchiolitis with an additional two cases showing bronchopneumonia. Multinuclear cells were seen in a subset of cases. In at least one case, small basophilic cytoplasmic inclusions and larger eosinophilic cytoplasmic inclusions were identified (Figure 1 and S1). No microthrombi were identified in the pulmonary system.

**Figure 1:**
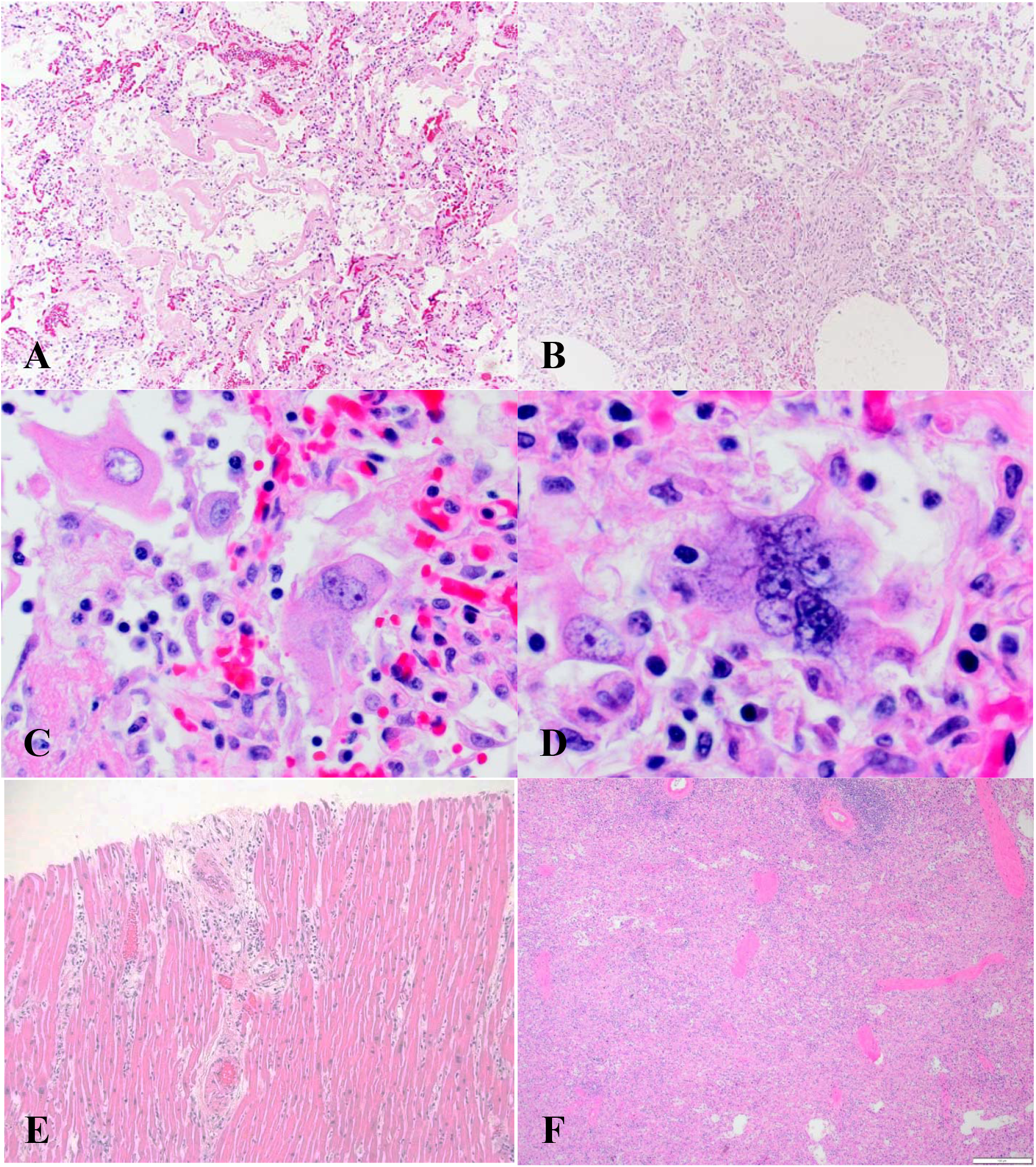
Histopathology of Fatal COVID-19 Infections. (A) Hyaline membranes. Hematoxylin and eosin; ×100. (B) Diffuse alveolar damage, organizing phase. Hematoxylin and eosin; ×100. (C) Multinucleated giant cells and pleomorphic, reactive pneumocytes. Hematoxylin and eosin; x400. (D) Multinucleated giant cells. Hematoxylin and eosin; x400. (E) Heart with lymphocytic myocarditis and associated myocyte damage. Hematoxylin and eosin; ×100. (F) Spleen with decreased white pulp. Hematoxylin and eosin; ×100.

Proximal and distal tracheal sections displayed mild to moderate edema; acute tracheitis was rarely observed. (Figure S2) The majority of cardiac findings were associated with prior injury or hypertensive changes; however, one case contained foci of lymphocytic inflammation associated with acute myocyte necrosis, consistent with lymphocytic myocarditis. Three cases showed evidence of splenic white pulp depletion. (Figure 1)

The kidneys showed chronic changes consistent with arterionephrosclerosis. One autopsy showed prominent proximal tubular epithelial cell vacuolization consistent with acute tubular injury. Autolysis precluded evaluation for acute tubular injury in other autopsies. Pathologic findings in the liver demonstrated chronic changes associated with pre-existing comorbidities. (Figure S2) Liver inflammation was not prominent, although some patients displayed mild periportal lymphocytic inflammation. Samples of thyroid, pituitary, adrenals, and pancreas were largely unremarkable. Post-mortem autolysis prevented a thorough investigation of the gastrointestinal system. Table 2 provides an overview of relevant histopathologic observations.

**Table 2:** Histologic Features of Patients with Fatal COIVD-19 Infections(n=12)

| **Pulmonary Findings** | **Number of patients** | **Cardiac findings** | **Number of patients** |
| --- | --- | --- | --- |
| Reactive pneumocytes | 12 (100%) | Myocyte hypertrophy | 12 (100%) |
| Pulmonary edema | 10 (83.3%) | Interstitial fibrosis | 10 (83.3%) |
| Diffuse alveolar damage* | 9 (75%) | Histologic evidence of remote infarct | 3 (25%) |
| Bronchopneumonia, neutrophilic bronchitis, or bronchiolitis | 8 (66.7%) | Amyloidosis | 1 (8.3%) |
| Organizing phase of diffuse alveolar damage | 6 (50%) | Lymphocytic myocarditis | 1 (8.3%) |
| Multinucleated giant cells | 4 (33.3%) | **Hepatic findings** | **Number of patients** |
| Background emphysematous changes | 4 (33.3%) | Congestion | 9 (75%) |
| Alveolar septal thickening | 2 (16.6%) | Steatosis | 8 (66.7%) |
| Intranuclear inclusions | 1 (8.3%) | Periportal lymphocytic inflammation | 3 (25%) |
| Microthrombi | 0 (0%) | Centrilobular necrosis | 2 (16.6%) |
| **Tracheal findings** | **Number of patients** | Lobular neutrophilic inflammation | 1 (8.3%) |
| Chronic (lymphocytic) tracheitis | 10 (83.3%) | **Gastrointestinal findings** | **Number of patients** |
| Edema | 9 (75%) | Multifocal gastric hemorrhage | 1 (8.3%) |
| Acute (neutrophilic) tracheitis | 4 (33.3%) | **Renal Findings** | **Number of patients** |
| Fibrosis or ossification | 1 (8.3%) | Mild to moderate arterionephrosclerosis | 9 (75%) |
| **Splenic findings** | **Number of patients** | Moderate to severe arterionephrosclerosis | 3 (25%) |
| Reduced white pulp | 3 (25%) | Diabetic changes | 3 (25%) |
| **Brain findings** | **Number of patients** | Amyloidosis | 1 (8.3%) |
| Punctate subarachnoid hemorrhages | 1 (8.3%) |  |  |
| Punctate brainstem hemorrhages | 1 (8.3%) |  |  |

\*Diffuse alveolar hemorrhage is defined by presence of hyaline membranes

### Electron Microscopy Findings

Electron micrographs demonstrate uniform, round viral particles ranging in size from ∼70-100 nm in the lung, trachea, kidney, and large intestines (Figure 2). Definitive viral particles were not observed in the other organs examined. The particles were found both individually or in aggregates in the cytoplasm or inside vesicles. In certain planes of section, the lipid membrane and the peripheral spikes (∼15 nm) were more clearly observed (Figure 2). Viral particles were present in both type I and type II pneumocytes. Type II pneumocytes showed numerous autophagosomes, characterized by double membranes and presence of organelles, in the cytoplasm. Some of these autophagosomes contained viral aggregates. No conclusive aggregates of nucleoprotein or viral inclusions were observed. Viral particles were also present in tracheal epithelial cells and within the extracellular mucus in the tracheal lumen. Within the kidney, viral particles localized to tubular epithelium, most prominently in the proximal tubules, and endothelial cells. Rare vesicle bound viral aggregates were also observed in the podocytes.

**Figure 2:**
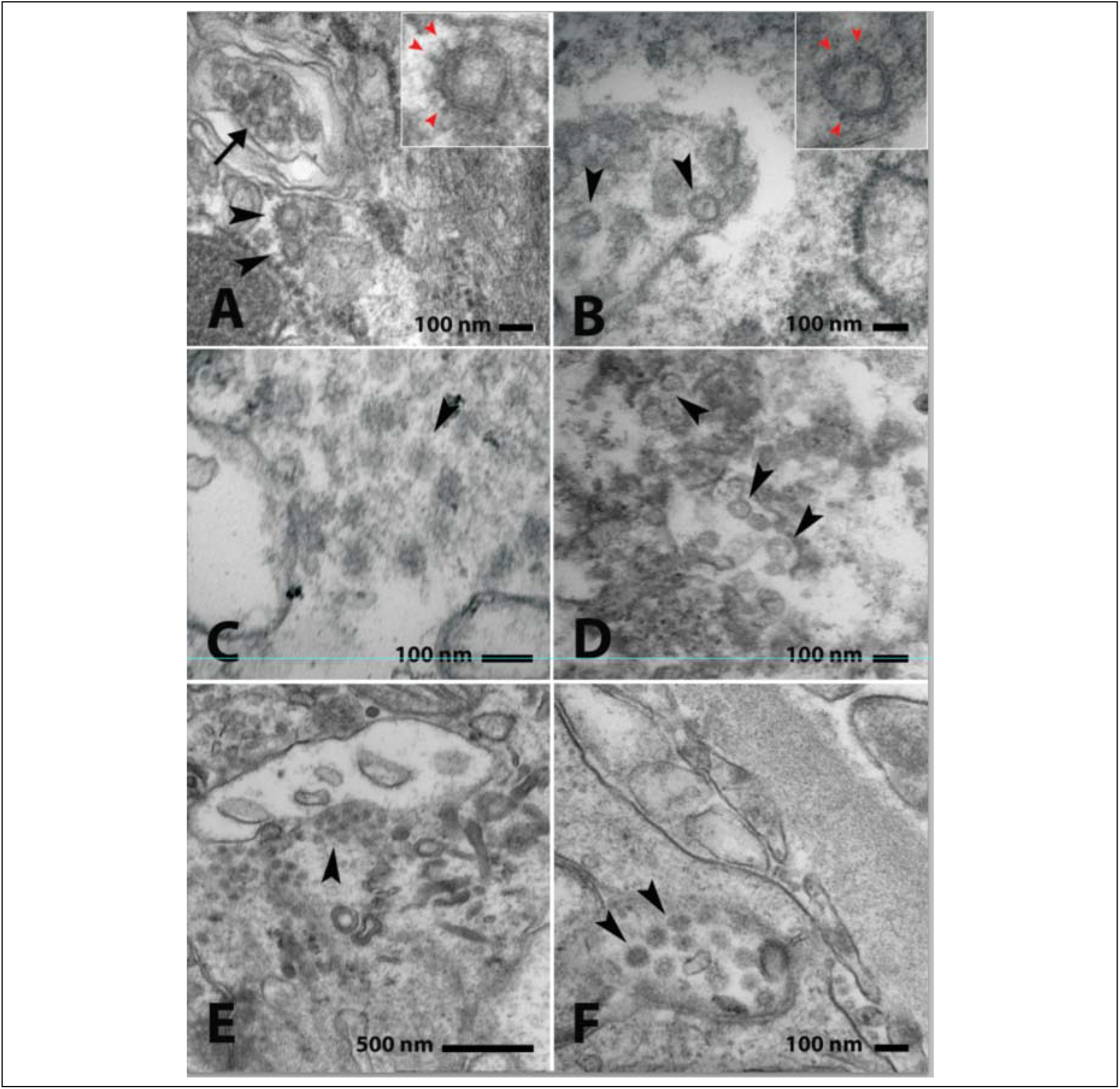
Ultrastructural Features in Fatal COVID-19 Infections. **(A) Lung, type II pneumocyte:** Viral particles inside a vesicle (black arrow) and in the cytoplasm (black arrowheads). The inset shows magnified view of one of the particles with peripheral spikes (red arrowheads). **(B) Trachea, epithelial cell:** Viral particles inside a vesicle (black arrowheads). The inset shows magnified view of one of the particles with peripheral spikes (red arrowheads). **(C) Trachea, extracellular mucus**: Viral particles (black arrowheads) mixed with cellular debris in tracheal lumen. **(D) Large intestine:** Viral particles (black arrowheads) in a degenerated mucosal epithelial cell. **(E) Kidney, proximal tubular epithelial cells:** Cytoplasmic viral particles aggregating in a cluster. **(F): Kidney, glomerular endothelial cell:** Viral particles within a vesicle with features consistent with an autophagosomes.

### Viral RNA Detection

Detectable levels of viral RNA were identified in multiple organs, most predominately the lungs (Table 3). Ct values ranged from 21 to 30 in respiratory tissues. Less abundant viral RNA was noted in the subcarinal lymph nodes (Ct=36). Other large organs including the kidney, heart, liver, spleen, and large intestines had RNA signals at Ct values greater than > 37. Viral RNA was not detected in the brain, bladder, esophagus, or stomach.

**Table 3:** Molecular and ultrastructural detection of SARS-CoV-2 in different tissue types.

| Sample | PCR (N1) | PCR (N2) | EM |
| --- | --- | --- | --- |
| Left lung | + | + | + |
| Right lung | + | + | + |
| Trachea (distal) | + | + | + |
| Trachea (proximal) | + | + | N/A |
| Lymph node | +/- | + | N/A |
| Large intestine | +/- | + | + |
| Heart | +/- | +/- | - |
| Liver | +/- | +/- | - |
| Spleen | +/- | +/- | - |
| Kidney | +/- | - | + |
| Small intestine | - | - | - |
| Brain | - | - | - |
| Bladder | - | - | N/A |
| Stomach | - | - | N/A |
| Esophagus | - | - | N/A |

Samples with Ct values < 40 were considered positive. Ct values $\leq 36$ noted with (+) and Ct values between 37 to 40 as (+/-). Tissues containing uniform, ~70-100nm enveloped particles with surface spikes were classified as positive (+) by EM.

## Discussion

Few reports have examined the histopathologic changes associated with SARS-CoV-2 infection and none have performed complete evaluation of all major organs.(7–9) Here, we present the largest summary of autopsy findings in patients who died following SARS-CoV-2 infection. Our results demonstrate the central role of lung damage in cases of terminal COVID-19 and we propose a mechanism for the extra-pulmonary spread of SARS-CoV-2.

Our cohort of patients exhibited similar comorbidities to those found in hospitalized COVID-19 patients described in other studies.(12) The most common conditions were cardiac disease, hypertension, diabetes, and obstructive sleep apnea. Patients with these conditions are often treated with angiotensin-converting enzyme inhibitors (ACEi) or angiotensin II type-1 receptor blockers. However, treatment with these therapies leads to upregulation of the ACE2 receptor which also serves as the cellular receptor for viral entry.(13) This observation has led to the hypothesis that upregulation of ACE2 may contribute to the high mortality rate in these patients.(14) Only two patients in our cohort had documented prescriptions for ACEi. ACE2 is expressed across a range of cell types and provides context to our observation of the virus in multiple different organ systems.(15)

The majority of the decedents presented with cough, shortness of breath, and fever which is similar to what has been described in other patients worldwide.(12) Upper respiratory symptoms such as sneezing, runny nose, and congestion were not common and highlights the predilection of SARS-CoV-2 for the lower respiratory tract as supported by our electron microscopy and PCR findings.

The most significant pathological findings in our cohort of fatal COVID-19 cases involved the pulmonary system. The majority of patients demonstrated pulmonary edema, pronounced reactive type II pneumocytes, intraalveolar fibrin, and hyaline membranes consistent with DAD. These findings are similar to those described during the 2002-2003 SARS (SARS-CoV) outbreak and more recent COVID-19 autopsies.(8,16,17) In contrast to SARS, multinucleated giant cells were seen in a minority of our cases.(17) We observed one case with cytoplasmic inclusions in the pneumocytes. The etiology of these inclusions in SARS-CoV-2 may be explained by in vitro ultrastructural studies of SARS-CoV where nucleoprotein aggregates were identified in the cytosol.(18)

Interestingly, the organizing phase of DAD was observed in patients with a brief symptom onset-to-death interval, which is in contrast to SARS where it was predominantly observed in those with longer hospitalizations.(19) As suggested by abnormal pulmonary CT scan findings in asymptomatic patients, the early changes of DAD may occur during the sub-clinical phase with symptom onset occurring in the organizing phase of damage.(20)

During the 2009 H1N1 pandemic, secondary bacterial pneumonia complicated approximately 25% of cases and was a significant cause of morbidity and mortality. (21) In our cohort, four patients had clinically documented secondary bacterial infection or histologic evidence of bronchopneumonia. This rate is much higher than what has been documented in other studies of patients admitted to the ICU.(22) Secondary bacterial infections may serve as a negative prognostic indicator for hospitalized patients.

Electron micrographic studies demonstrated infection of type I and type II pneumocytes by virus, which recapitulates findings documented in SARS-CoV.(16) We did not observe definitive involvement of alveolar macrophages, though this may be the result of limited sample size and post-mortem degeneration.(23) We also observed the induction of double-membrane vesicles within infected pneumocytes, consistent with autophagosomes. Studies of the SARS-CoV replication pathway raised the hypothesis that these double-membrane vesicles served to shield the virus from host immune detection.(24)

Our RNA data and electron microscopy images suggest the infectious burden of SARS-CoV-2 centers in the lower respiratory tree, specifically the alveolar epithelium. Sites higher in the respiratory tree had virus detected but at a lower relative concentration compared to alveolar tissue. These findings could represent contamination by alveolar secretions or decreased susceptibility of the upper airway to viral infection. We favor the latter interpretation as ISH/IHC studies of SARS patients demonstrated reduced, but detectable SARS-CoV in the bronchioles.(25) Moreover, it is likely that high expression of ACE2 in type II pneumocytes makes these cells more susceptible to SARS-CoV-2 infection.(26,27)

Detection of moderate levels of viral RNA in the sub-carinal lymph node suggests leukocytes may serve as a route for the virus to disseminate from the alveoli to other organs. While fulminant infection of other organs may not occur, given the pre-existing comorbidities in our patient population, we hypothesize that even a low level of virally-induced cytotoxicity could prompt organ dysfunction. Infection of leukocytes also raises the question as to the degree which SARS-CoV-2 infection modulates the immune response and contributes to cytokine dysregulation. This hypothesis is in keeping with the pathologic mechanism postulated for SARS.(28)

Our understanding of the cardiac effects of SARS-CoV-2 infection are rapidly evolving. In a study documenting the clinical course of ICU patients in Kirkland, Washington, it was found that 33% of patients experienced cardiomyopathy of unclear etiology.(29) Similarly, myocardial injury with elevated troponins was identified in 22% of ICU patients.(12) While the majority of our patient cohort had no clinical concern for cardiomyopathy, one case from a patient with elevated troponins demonstrated lymphocytic myocarditis with detectable SARS-CoV-2 RNA.

The kidneys demonstrated variable chronic changes such as arterionephrosclerosis and diabetic nephropathy in keeping with the poor renal function of our cohort. No specific viral cytopathic changes or increased inflammation were identified by light microscopy. However, by electron microscopy, we identified abundant viral particles in proximal tubules, relatively fewer in the endothelial cells, and rare particles in the podocytes. High expression of ACE2 has recently been confirmed in proximal tubular epithelial cells.(26,27) The lack of inflammation in the kidneys contrasts with our pulmonary findings and raises the possibility of organ-specific viral responses and patterns of damage. Our findings have implications for transplant surgeries, as a negative nasopharyngeal swab may not exclude virus in other organs. These findings also highlight the peculiarity that virus has not been identified in the urine.(5)

Hepatic findings in our case series are similar to prior reports, however it is unclear if these changes are due to COVID-19 or a combination of background metabolic disease and general shock physiology.(9) The spleens in our cohort were largely unremarkable with a subset demonstrating white pulp depletion and one case of splenomegaly. Splenic white pulp depletion was also observed in SARS-CoV infections, but was associated with decreased spleen weight.(16) Virus observed in the lumen of the bowel is in keeping with studies demonstrating shedding in the stool.(30)

We acknowledge the inconsistency between our tissue PCR data and EM images for certain organs. In particular, abundant viral particles were observed in the kidney, however, only a weak RNA signal was detected. A potential explanation for this result may be due to sampling bias if virus distribution is patchy. Another cause for the low PCR signal (Ct > 37) detected across multiple organs could be related to post-mortem tissue autolysis and RNA degradation. Application of more sensitive technologies such as high-throughput sequencing or in situ hybridization will improve our ability to detect small fragments of RNA.

Our findings demonstrate the central role of the lower respiratory tree as the likely nidus from which virus disseminates to other organs. We found broad tropism for SARS-CoV-2 with virus identified in the kidneys, heart, large intestines, spleen, and liver. Accurate and comprehensive studies of the histopathologic manifestations of COVID-19 will guide clinicians and researchers towards reducing mortality with timely clinical and therapeutic interventions.

## Data Availability

All data will be made available by request to the corresponding author.

## Contributors

B.T.B. conceived and designed the study.

H.M., M.J., and I.C. contributed to clinical data collection.

H.M., B.N., and B.T.B. contributed to figure design.

N.Y., T.W., J.M.L., D.M, R.J., I.C., and B.T.B. performed the autopsies.

N.Y., T.W., J.M.L., D.M, R.J., I.C., H.M, H. X., B.N., and B.T.B. contributed to histopathologic evaluation of tissue.

B.N. and B.T.B. contributed to electron microscopy images.

B.T.B. and S.F. contributed to molecular testing.

B.T.B. and H.M. wrote the manuscript.

All authors contributed to data analysis, data interpretation, and editing the manuscript.

## Acknowledgments

We thank the technical staff of King County Medical Examiner’s Office and Snohomish Medical Examiner’s Office for their assistance in autopsy procedures.

We thank Ms. Jennifer Swicord, Xiaobing Ren and Mr. Gianni Niolu for preparation of electron microscopy specimens.

---

**Supplemental Figure 1:**
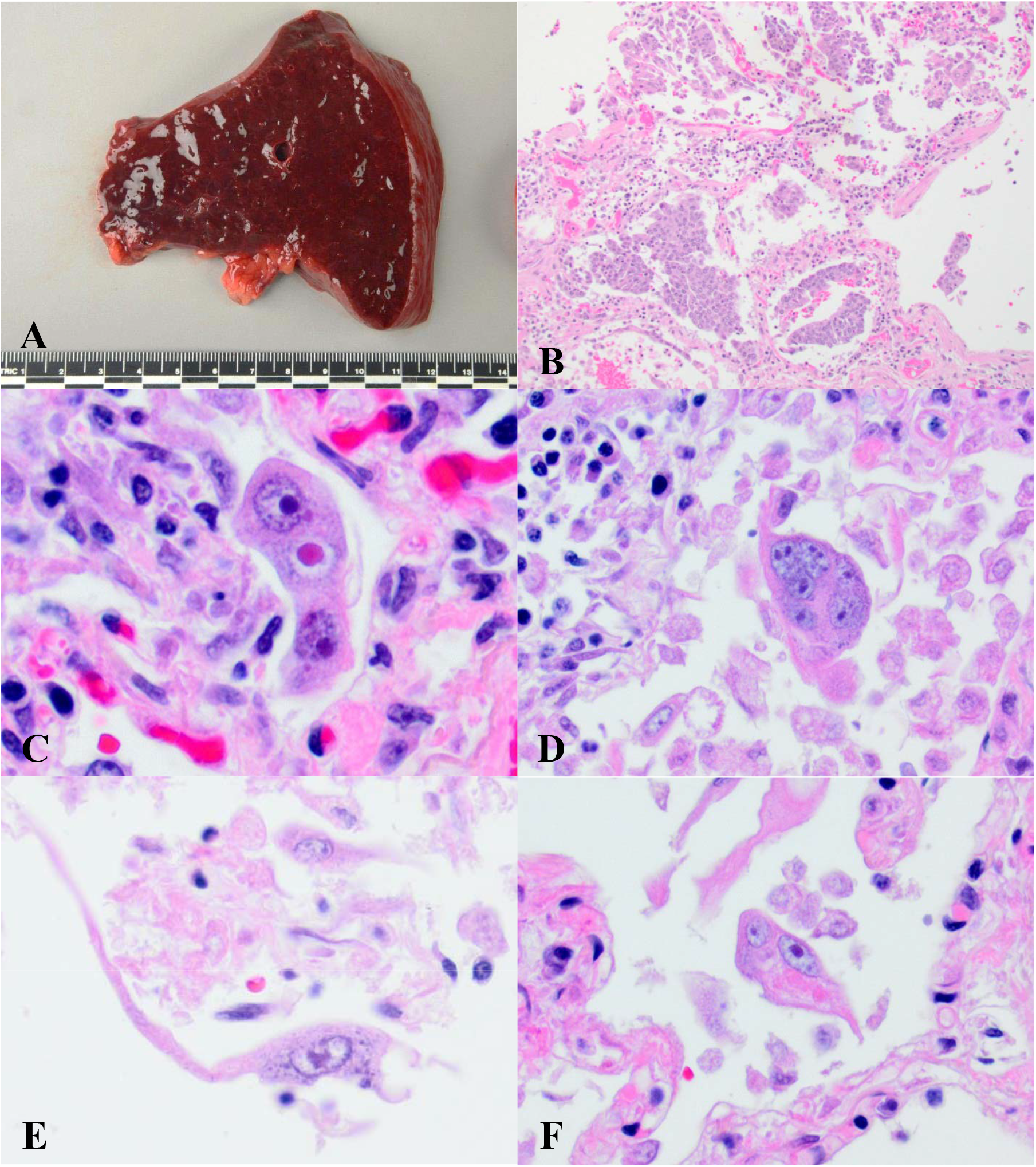
Additional Pulmonary Histopathology from Fatal COVID-19 Infections. (A) Edematous, heavy lung (B) Squamous metaplasia, Hematoxylin and eosin; ×100. (C) Reactive cell with hemophagocytosis. Hematoxylin and eosin; x400. (D) Multinucleated giant cell. Hematoxylin and eosin; x400. (E) Basophilic intracytoplasmic inclusions. Hematoxylin and eosin; x400. (F) Binucleated cell with amorphilic, eosinophilic cytoplasmic inclusion. Hematoxylin and eosin; x400.

**Supplemental Figure 2:**
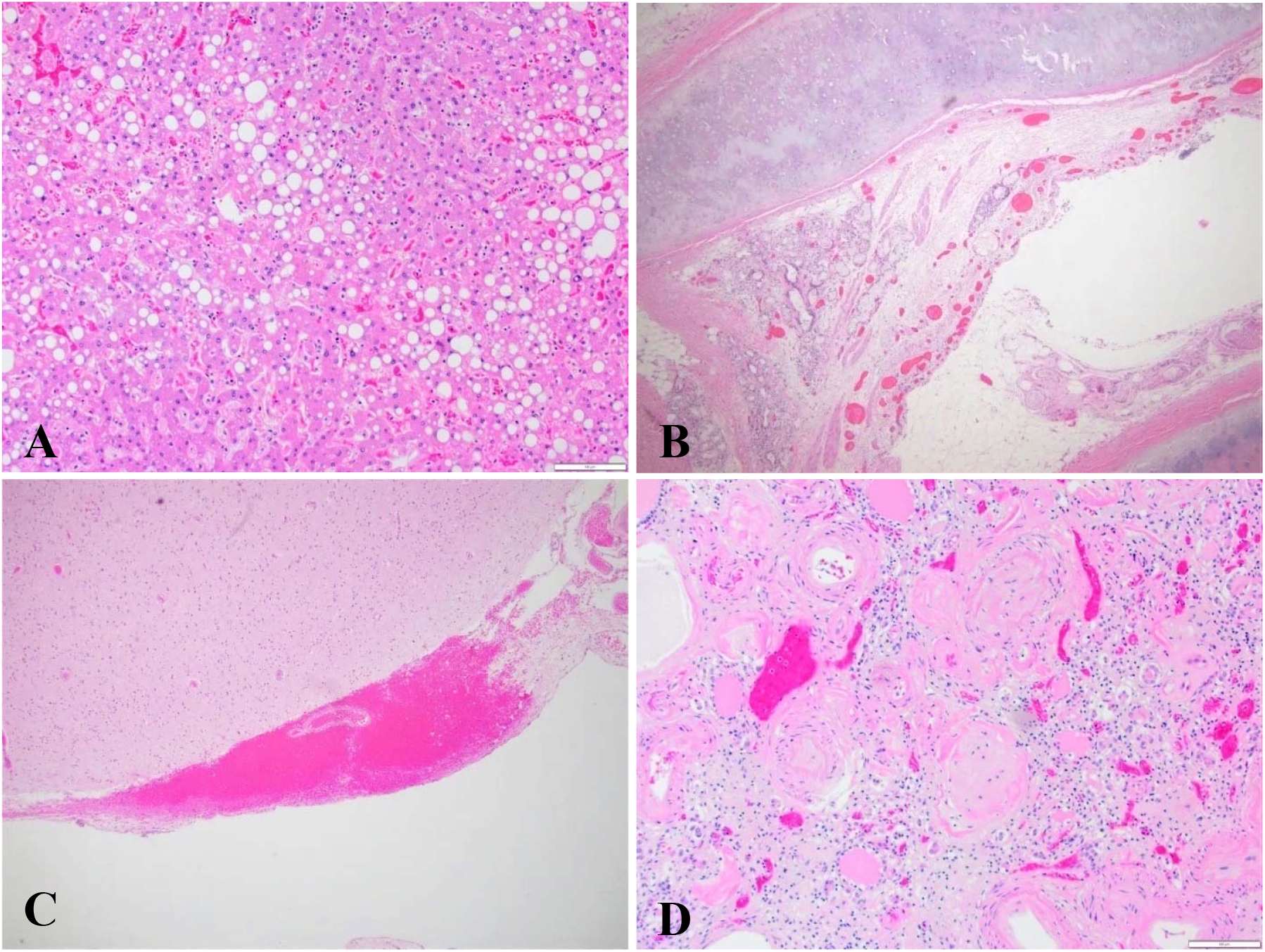
Histologic Findings of Major Organs in Fatal COVID-19 Infections. (A) Mixed micro and macrovesicular steatosis. Hematoxylin and eosin; ×100. (B) Trachea with congestion and edema. Hematoxylin and eosin; x40. (C) Brain with small, subarachnoid hemorrhage. Hematoxylin and eosin; ×100. (D) Kidney with severe arterionephrosclerosis. Hematoxylin and eosin; ×100.

**Supplemental Table 1:** Selected features of Patients with Fatal COVID-19 Infection (n=12)

|  |  |
| --- | --- |
| Median age (range) | 70.4 years (42-84) |
| Sex (Male: Female) | 5:7 |
| Median days from symptom onset to death (range) | 7 (1-14) |
| Number with antemortem SARS-CoV-2 confirmation | 10 (83.3%) |
| Number with post-mortem SARS-CoV-2 confirmation | 2 (16.7%) |
| Number of patients ACEi or AR2 blocker | 2 (16.7%) |
| Elevated liver enzymes | 1 (8.3%) |
| Leukocytosis | 2 (16.7%) |
| Leukopenia | 7 (58.3%) |
| Antemortem confirmation of secondary pulmonary infection | 2 (16.7%) |

Elevated liver enzymes defined by AST ≥ 109 U/l and ALT ≥ 97 U/l, leukocytosis defined by a white count over 11,000/ $\mu$ L, leukopenia defined by a white count under 4,000/ $\mu$ L. ACEi (angiotensin converting enzyme inhibitor), AR2 (angiotensin receptor 2).

**Supplemental Table 2:** Select Gross Findings in Fatal COVID-19 Infections.

| **Organ and findings** | **Average Weight (Grams) (n=5)** | **Number of Patients with Gross finding** |
| --- | --- | --- |
| **Right Lung /Left Lung** | 1049/755 |  |
| Edema |  | 5 (100%) |
| Hemorrhage |  | 1 (20%) |
| Consolidation |  | 0 (0%) |
| Pleural fluid (mL)* | 262/250 | 2 |
| **Heart** | 492 (range 393-685) |  |
| Cardiomegaly** |  | 3 (60%) |
| Coronary Artery Disease |  | 3 (60%) |
| Evidence of remote infarction |  | 0 (0%) |
| Evidence of acute infarction |  | 0 (0%) |
| **Liver** | 1749 (range 1068-2425) |  |
| Congestion |  | 5 (100%) |
| Cholestasis |  | 0 (0%) |
| Cirrhosis |  | 0 (0%) |
| **Spleen** | 209 (range 154-350) |  |
| **Right Kidney /Left Kidney** | 157/156 |  |
| Granular surface/scarring |  | 4 (80%) |
| Poor cortico-medullary differentiation |  | 1 (20%) |
| **Brain (n=4)** | 1395 |  |
| Meningeal punctate subarachnoid hemorrhages |  | 1 (20%) |

\* n = 2, \*\* cardiomegaly defined as a heart weight >450g,

